# Integrative analysis of viral entry networks and clinical outcomes identifies a protective role for spironolactone in severe COVID-19

**DOI:** 10.1101/2022.07.02.22277181

**Authors:** Henry Cousins, Adrienne Sarah Kline, Chengkun Wang, Yuanhao Qu, Mengdi Wang, Russ Altman, Yuan Luo, Le Cong

**Affiliations:** Department of Biomedical Data Science, Stanford University School of Medicine, Stanford, CA, USA; Medical Scientist Training Program, Stanford University School of Medicine, Stanford, CA, USA; Department of Preventive Medicine, Feinberg School of Medicine, Northwestern University, Chicago, IL, USA; Department of Pathology, Stanford University School of Medicine, Stanford, CA, USA; Department of Genetics, Stanford University School of Medicine, Stanford, CA, USA; Department of Electronic Engineering, Princeton University, Princeton, NJ, USA; Department of Medicine, Stanford University School of Medicine, Stanford, CA, USA; Department of Bioengineering, Stanford University, Stanford, CA, USA

**Author notes:** Correspondence to (H.C.), (R.A.), (Y.L.), (L.C.).

## Abstract

Treatment strategies that target host entry factors have proven an effective means of impeding viral entry in HIV and may be more robust to viral evolution than drugs targeting viral proteins directly. High-throughput functional screens provide an unbiased means of identifying genes that influence the infection of host cells, while retrospective cohort analysis can measure the real-world, clinical potential of repurposing existing therapeutics as antiviral treatments. Here, we combine these two powerful methods to identify drugs that alter the clinical course of COVID-19 by targeting host entry factors. We demonstrate that integrative analysis of genome-wide CRISPR screening datasets enables network-based prioritization of drugs modulating viral entry, and we identify three common medications (spironolactone, quetiapine, and carvedilol) based on their network proximity to putative host factors. To understand the drugs’ real-world impact, we perform a propensity-score-matched, retrospective cohort study of 64,349 COVID-19 patients and show that spironolactone use is associated with improved clinical prognosis, measured by both ICU admission and mechanical ventilation rates. Finally, we show that spironolactone exerts a dose-dependent inhibitory effect on viral entry in a human lung epithelial cell line. Our results suggest that spironolactone may improve clinical outcomes in COVID-19 through tissue-dependent inhibition of viral entry. Our work further provides a potential approach to integrate functional genomics with real-world evidence for drug repurposing.

## INTRODUCTION

Host cell entry represents a critical stage of the SARS-CoV-2 replication cycle that determines the tropism and virulence of emerging variants (1). SARS-CoV-2 entry relies canonically on binding between viral Spike protein and host ACE2, followed by processing of the Spike protein by endogenous proteases, most notably TMPRSS2 and furin (2, 3). However, the complete entry process relies both directly and indirectly on a network of hundreds of host genes that remains poorly understood (4, 5).

Heterogeneity in expression patterns of accessory entry factors, which facilitate viral adhesion, cleavage events, and membrane fusion, is a chief determinant of viral susceptibility in terms of both tissue types and patient subgroups (6, 7). Consequently, drug interactions with such host factors can promote or inhibit viral entry *in vitro* and have in some cases demonstrated clinical efficacy in large-scale studies (8–10). With the emergence of SARS-CoV-2 variants with significant genome-wide mutational load, such as the BA.2 and XE strains, therapeutics and vaccines targeting viral proteins of early strains have shown reduced efficacy in recent outbreaks (11–13). Further, structural and functional study of the Omicron mutational landscape revealed a range of adaptive mutations that facilitate antibody neutralization, vaccine protection, and high-affinity viral interactions with host ACE2 receptor (12, 14, 15). There consequently exists a pressing need to identify therapies that modulate host entry factors, which may be both more robust to future variants and complementary to existing direct-acting antivirals such as nirmatrelvir and molnupiravir (16, 17).

High-throughput forward genetic screens, primarily performed with CRISPR-Cas9 systems in knock-out (CRISPR-KO) and activation (CRISPRa) formats, provide a powerful tool to identify host genes that facilitate viral entry (18–25). Such methods enable causal inference of single-gene effects that may be confounded in gene expression assays by both epistatic patterns and immune mechanisms distinct from viral entry. CRISPR screens can also quantify gene effects in distinct cell types and different perturbation schemas, which provides specific mechanistic insights but can limit the generalizability of findings from any individual experiment (26).

We hypothesized that integrated analysis of multiple viral-entry functional screens would reveal a shared network of host entry genes, with more generalizable implications for drug repurposing than would be possible using individual datasets. We performed drug-target network analysis using all publicly available, genome-wide CRISPR screens of SARS-CoV-2 viral entry, which identified three common drugs, spironolactone, carvedilol, and quetiapine, as potential modulators of viral entry. Furthermore, we conducted a retrospective clinical outcome analysis of these drugs using medical records from 64,349 COVID-19 patients, which supported a significant protective role for spironolactone. Finally, we demonstrated that spironolactone exerts a time-dependent inhibitory effect on SARS-CoV-2 viral entry in human lung cells, suggesting that spironolactone may mediate a milder disease course by suppressing viral entry.

## METHODS

### CRISPR screening data

We identified all published, genome-wide CRISPR-KO screens of SARS-CoV-2 entry in human cells available as of May 2022, with five screens meeting criteria. Three screens were performed in HuH-7 liver cells, and two were performed in A549 lung epithelial cells. Multiplicities of infection (MOI) for each screen ranged from 0.21 to 0.4. We also identified all genome-wide CRISPRa screens in human cells available as of May 2022, with three screens meeting criteria. These included two screens in Calu-3 lung epithelial cells and one in HEK293T embryonic kidney cells, with MOI ranging from 0.05 to 0.5. Raw data for each screen was obtained from the respective source publication (Figure 1B). For each screen, fold-change values were converted to Z-scores, with positive scores indicating genes whose expression promoted viral entry.

**Figure 1.**
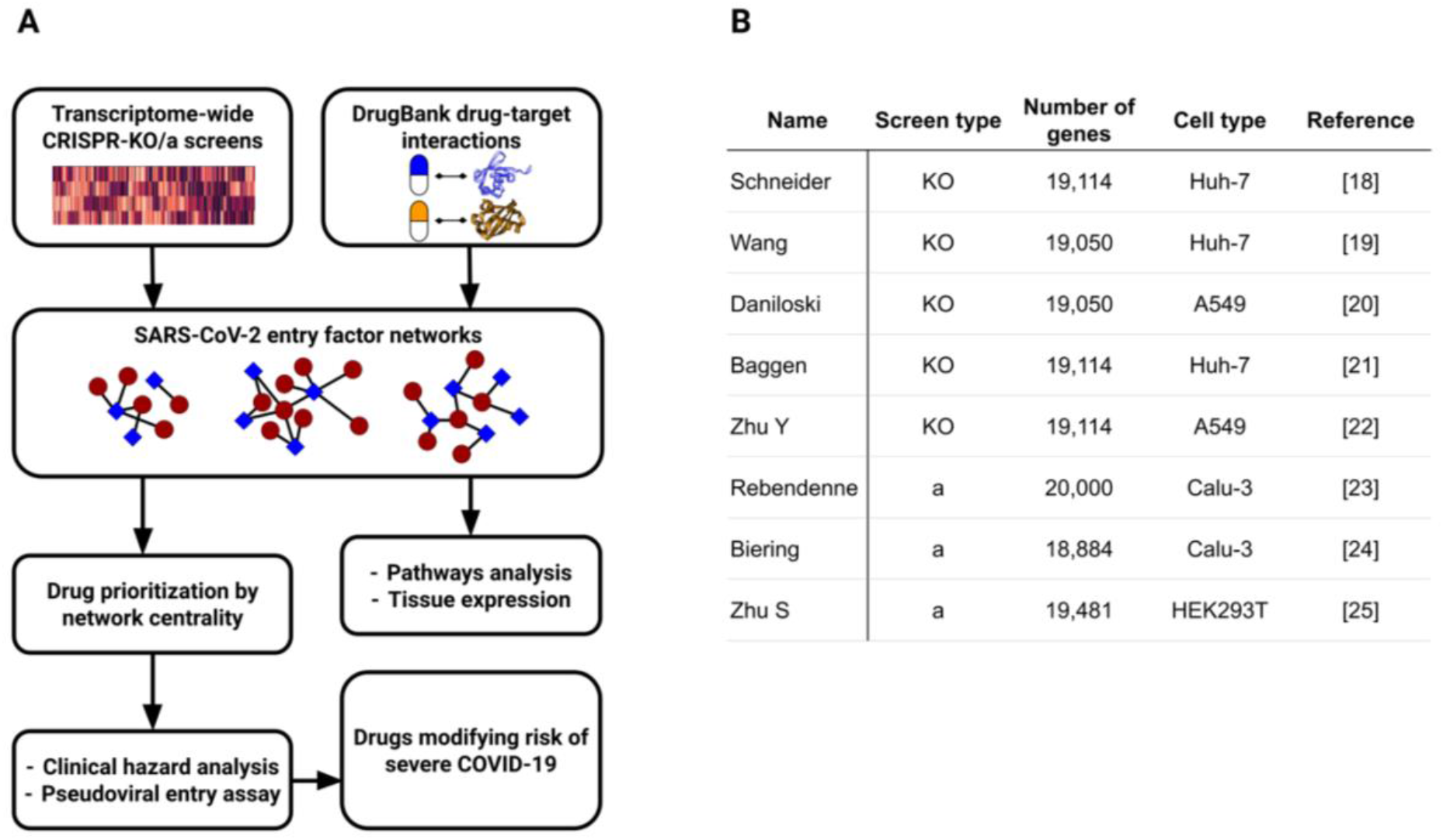
Integrated analysis of SARS-CoV-2 entry networks for drug repositioning. A. Genome-wide gene essentiality scores for viral entry were obtained from five CRISPR-KO screens and three CRISPRa screens. These were combined with high-confidence drug-target interactions from DrugBank to generate drug-gene subnetworks for hit genes from each screen. Drugs are prioritized based on normalized degree centrality within each subnetwork, and top drugs are validated through propensity-score matched hazard analysis of retrospective electronic medical record data. B. Descriptions of each functional screen considered in the analysis.

### Functional enrichment analysis

Enrichment analyses were performed using the prerank gene set enrichment analysis (GSEA) algorithm. KEGG pathway gene sets were obtained from the KEGG Pathway Database, and gene ontology molecular function (GOMF) gene sets were downloaded from the MSigDB Signatures Database (27, 28). In all cases, sets with a minimum size of 3 and maximum set size of 500 were considered. We defined significance as a false discovery rate (FDR) less than 0.05.

### Network-based drug rankings

Drug-gene interactions for FDA-approved drugs were obtained from DrugBank and used to construct a bipartite graph for each screen containing two node classes, compounds and genes, and undirected edges representing known interactions between protein-coding genes and FDA-approved drugs (29). The complete network contained 7,233 nodes (4,927 compounds and 2,306 genes) and 14,863 edges. For each screen, a unique subgraph was defined encompassing gene hits, measured as the top 5% of genes by Z-score, and their corresponding drug interactions.

In each subgraph, compounds were ranked by normalized degree centrality (NDC), defined as node degree normalized to the total number of possible neighbors, reflecting each compound’s proximity to host entry factors identified in the screen. Alternative measures of centrality, including betweenness centrality and eigenvector centrality, were also evaluated. We defined drug hits as the top 1% of drugs by NDC rank for each screen and selected for follow-up all drugs identified as hits in at least three screens.

### Cohort construction

De-identified patient medical records were obtained from the Northwestern Medicine Electronic Data Warehouse. Records were filtered to include only patients who had a positive COVID-19 test by means of reverse transcription-polymerase chain reaction (RT-PCR) testing. Patients who had more than one positive test were sorted by date, and the first was selected to create a unique set of patient identifiers. A 6-month time window preceding the first positive COVID-19 test was created for use as a filter for medication treatment status.

Patient characteristics that served as covariates considered in the analysis included race, gender, age, postal code, and medical comorbidities such as hypertension, diabetes, metastatic cancer, rheumatological disease, autoimmune disorders, and kidney failure, which are reported according to Charlson Comorbidities (30). The full list of comorbidities is provided as supplementary material (**Supplementary Table S1**). Records in which both race and postal code were missing (n = 8,201) were removed, as data was not missing at random. The final database included 64,349 unique patients with a positive COVID-19 test and complete comorbidity labels.

For each of the medications under investigation, treatment groups were identified by string matching to generic/brand names for the medication of interest. We defined users as those with medication orders occurring within 6 months preceding their first recorded COVID-19-positive specimen, such that treatment status connotes drug exposure prior to COVID-19 diagnosis.

### Propensity score matching

Propensity score matching was performed using the *psmpy* package in Python 3.7 (31). In our use case, logistic regression was executed where the treatment state (for the medication of interest) was regressed on the set of covariates defined previously. Due to the size imbalance between treatment and control group sizes, logistic regression was run repeatedly on a balanced sample to generate a probability integer for each observation by folding iteratively over the larger class and averaging over repeated patient indexes. The logit of the logistic regression prediction was calculated for control and treatment observations, as it more closely approximates a normal distribution (32). Following this, a unidimensional k-nearest neighbors (k-NN) algorithm was fitted to the logit scores of the control group. The treatment group was then fitted to the model, calculating Euclidean distance as the similarity metric. Am exclusionary caliper size of 0.25 of the standard deviation of the distance was implemented to reduce distant matches. To prevent samples from sharing the closest first match, matching was performed without replacement. In this way, treatment-control patient pairs were identified in which each subject had an approximate equal likelihood of receiving the drug of interest.

To verify adequate matching, a Cohen’s D statistic (standardized mean difference) was calculated before and after matching, ensuring that the matched cohort had smaller covariate effect sizes. Additionally, the two cohorts’ logit scores and the number of patients in each category were compared to confirm similarity.

### Statistical analysis

Clinical outcomes under investigation included admission to the intensive care unit (ICU) and initiation of mechanical ventilation. Odds ratios for each clinical endpoint, with corresponding confidence intervals and p-values, were calculated using both McNemar’s test for matched treatment-control pairs and a chi-square test for the same cohorts without pairing.

### Ethics statement

The clinical study was approved by the Northwestern University Institutional Review Board.

### Software

All computational analyses were performed in a Python 3.7 environment.

### Replicating vesicular stomatitis virus (VSV) pseudovirus generation

Recombinant VSV expressing eGFP in the 1st position (VSVdG-GFP-CoV2-S) was generated using plasmid-based methods. The plasmid to rescue this virus was generated by inserting a codon optimized SARS-CoV2-S based on the Wuhan-Hu-1 isolate (Genbank:MN908947.3), which was mutated to remove a putative ER retention domain (K1269A and H1271A) into a VSV-eGFP-dG vector (Addgene, Plasmid #31842) in frame with the deleted VSV-G. The control virus VSVdG-RABV-G SAD-B19 was also generated by inserting Rabies virus G in the same vector. Both viruses were rescued in 293FT/VeroE6 cell co-culture and amplified in VeroE6 cells and titrated in VeroE6 cells over-expressing TMPRSS2. Sequencing of the amplified virus revealed an early C-terminal Stop signal (1274STOP) and a partial mutation at A372T (∼50%) in the ectodomain.

### Pseudovirus infection assay using Replicating VSV pseudovirus

HEK293FT cells were plated in clear 96-well plates at 2×104 cells per well approximately 24 hours prior to infection in 100 uL of media containing 10% FBS. Cells were infected with VSVdG-CoV2-S or VSVdG-RABV-G at an MOI of 0.1. Infection was performed by diluting virus in media without FBS and adding 150 uL of diluted virus per well. After addition of the virus, the plate was spun at 900 x g for 60 minutes at 30°C. Infection was tracked over time using an Incucyte system (Sartorius) in a 37°C and 5% CO2 incubator using 4x magnification and detecting GFP. GFP+ cells were counted using Incucyte Analysis software and data were reported as GFP positive foci per well after normalization to confluence.

### In vitro viral entry inhibition experiments

All inhibitor assays use 96-well plates coated with Poly-D-Lysine (Thermo Fisher, A3890401) at a concentration of 50 ug/mL for 2 hours at room temperature. The plates were then washed with PBS three times, and 1 × 10^4^ cells were plated in a final volume of 100 uL of culture media. The next day, 20 uL of media was removed from each well and replaced with a 5X concentration of the inhibitor in culture media at the indicated dilution. The cells were then returned to 37°C. Two hours later, diluted SARS-CoV-2 Spike pseudotyped lentiviruses (for an ∼MOI of 0.05-0.15) were added to each well. Plates were spinfected and assayed as described above. Drugs (Sigma-Aldrich) were diluted in PBS via vigorously vortexing to a concentration of 100 mM prior to dilution in culture media.

## RESULTS

To identify host subnetworks that facilitate SARS-CoV-2 viral entry, we obtained all genome-wide CRISPR screens measuring the impact of individual genes’ expression on viral infection in human cells (**Figure 1A**). The screens accounted for a variety of cellular contexts, including both lung and non-lung cell types, and functional perturbations, namely loss-of-function (i.e., CRISPR-KO) and gain-of-function (i.e. CRISPRa) screens (**Figure 1B**). The final dataset collection included five CRISPR-KO and three CRISPRa screens, enabling resolution of context-dependent entry mechanisms that could not be identified by only the KO datasets.

The eight screens exhibited variable levels of correlation at the single-gene level, consistent with their heterogeneous cellular contexts (**Figure 2A**). 88% (7/8) of screens were significantly correlated with at least one other screen, while 26% (7/28) of all pairwise comparisons revealed significant positive correlation after correcting for multiple testing (**Figure 2B**). Gene-level agreement was higher within each class of screen, with significant positive correlation among 60% (6/10) of CRISPR-KO and 33% (1/3) of CRISPRa screen pairs. Both pairs of screens performed in lung-derived tissue were significantly positively correlated at the single gene level, but cellular context was not significantly associated with gene-level correlation overall (p = 0.0825).

**Figure 2.**
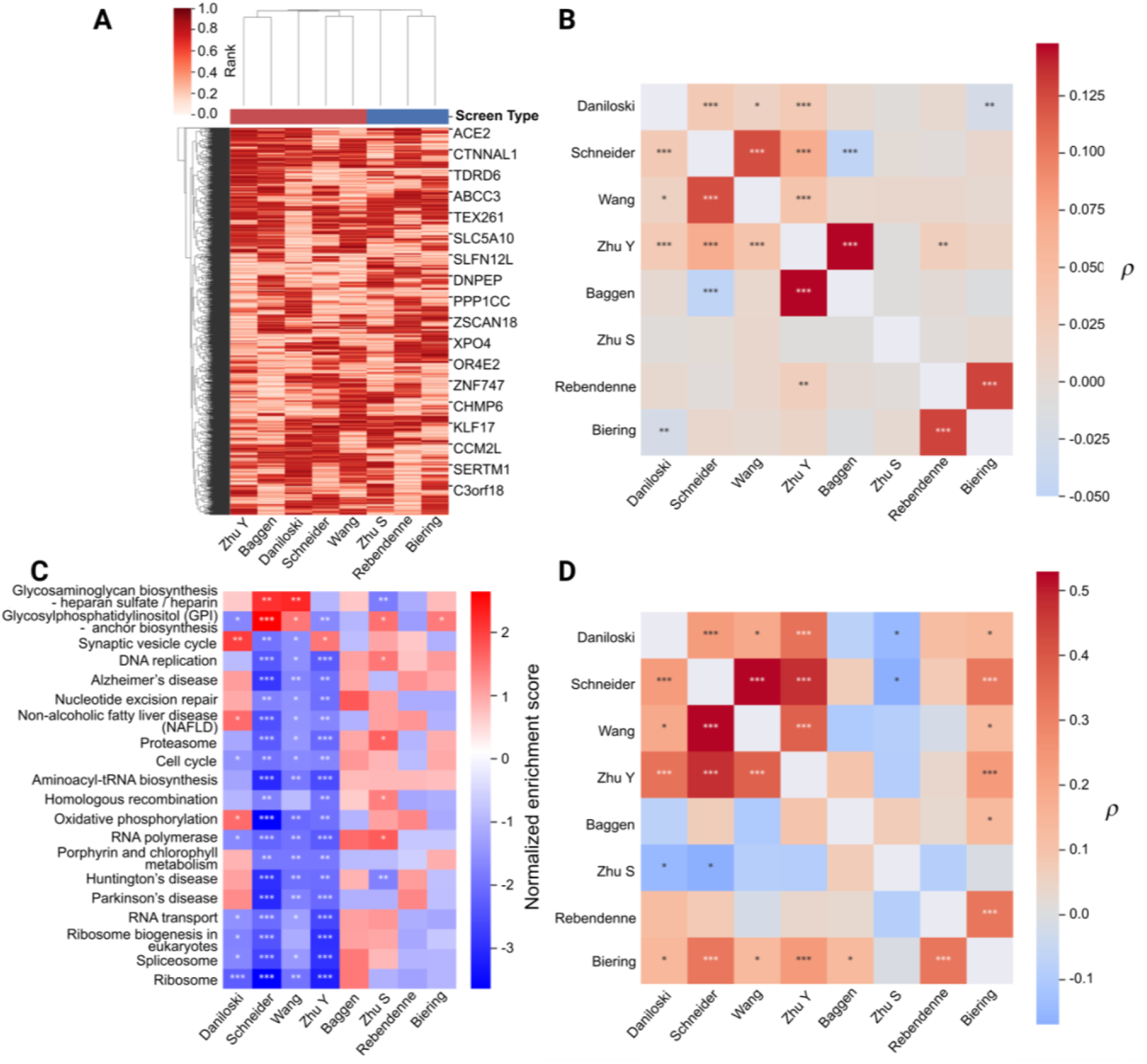
Correlations and functional enrichment patterns among screens. A. Cluster map of individual gene ranks for all screens. Red bars indicate knockdown screens, and blue bars indicate activation screens. Hierarchical clustering groups knockdown and activations screens together. B. Pairwise correlation measurements among screens. Annotations indicate significance at the nominal level (*), after correcting for multiple testing among screens (**), and after correcting for multiple testing among all pairs (***). A majority of knockdown screens demonstrate correlation at a nominal or higher significance. Correlations between knockdown and activation screens exist at a lower frequency. C. Pathways with significant enrichment in multiple screens involve cellular adhesion and synaptic transport. Annotations indicate significance at FDR values of 0.4 (*), 0.05 (**), and 0.001 (***). D. Inter-screen correlations for KEGG pathway enrichment scores are stronger than for individual gene scores. Annotations are the same as in (B).

We next quantified functional pathway enrichment among each screen using GSEA. 20 KEGG pathways were significantly enriched in at least two screens, including several pathways with known involvement in SARS-CoV-2 entry (**Figure 2C**). Pathways involved in glycosaminoglycan and phosphoglycerides were most strongly enriched, consistent with their essential role in viral attachment (33). We also observed significant de-enrichment of pathways involved in neurodegenerative disease, including Alzheimer’s, Huntington’s, and Parkinson’s diseases, as well as synaptic signaling broadly. Correlations among normalized pathway enrichment scores for each screen were generally higher than correlations for individual genes, although screens with higher gene-level correlations tended to have higher pathway correlations as well (**Figure 2D**).

We next constructed unweighted networks representing known interactions between FDA-approved drugs and entry genes identified in individual screens (**Figure 3A**). Each network contained an average of 116 (standard deviation 8.00) genes, 605 (208) drugs, and 758 (328) edges each, corresponding to an average density of 1.06% (0.112%). The mean degree of each graph was 6.50 (2.86) for gene nodes, 1.23 (0.104) for drug nodes, and 2.03 (0.256) overall, and 3.81% (1.45%) genes per screen, on average, lacked known interactions with any drugs. We prioritized drugs for downstream analysis based on centrality within each hit network. As both local (NDC) and global (betweenness) measures of centrality yielded similar rankings (*ρ* = 0.99, p < 0.001), we used NDC for subsequent analyses due to its interpretability. No drug met significance in all screens, while 248 drugs were significant in at least one dataset. 33 drugs met significance in at least three datasets (**Figure 3B**). Drug hits encompassed a range of functional categories, with a predominance of psychoactive compounds. Tricyclic antidepressants were the most common category, comprising 18.2% (6/33) of hits, followed by atypical antipsychotics, which comprised 9.1% (3/33).

**Figure 3.**
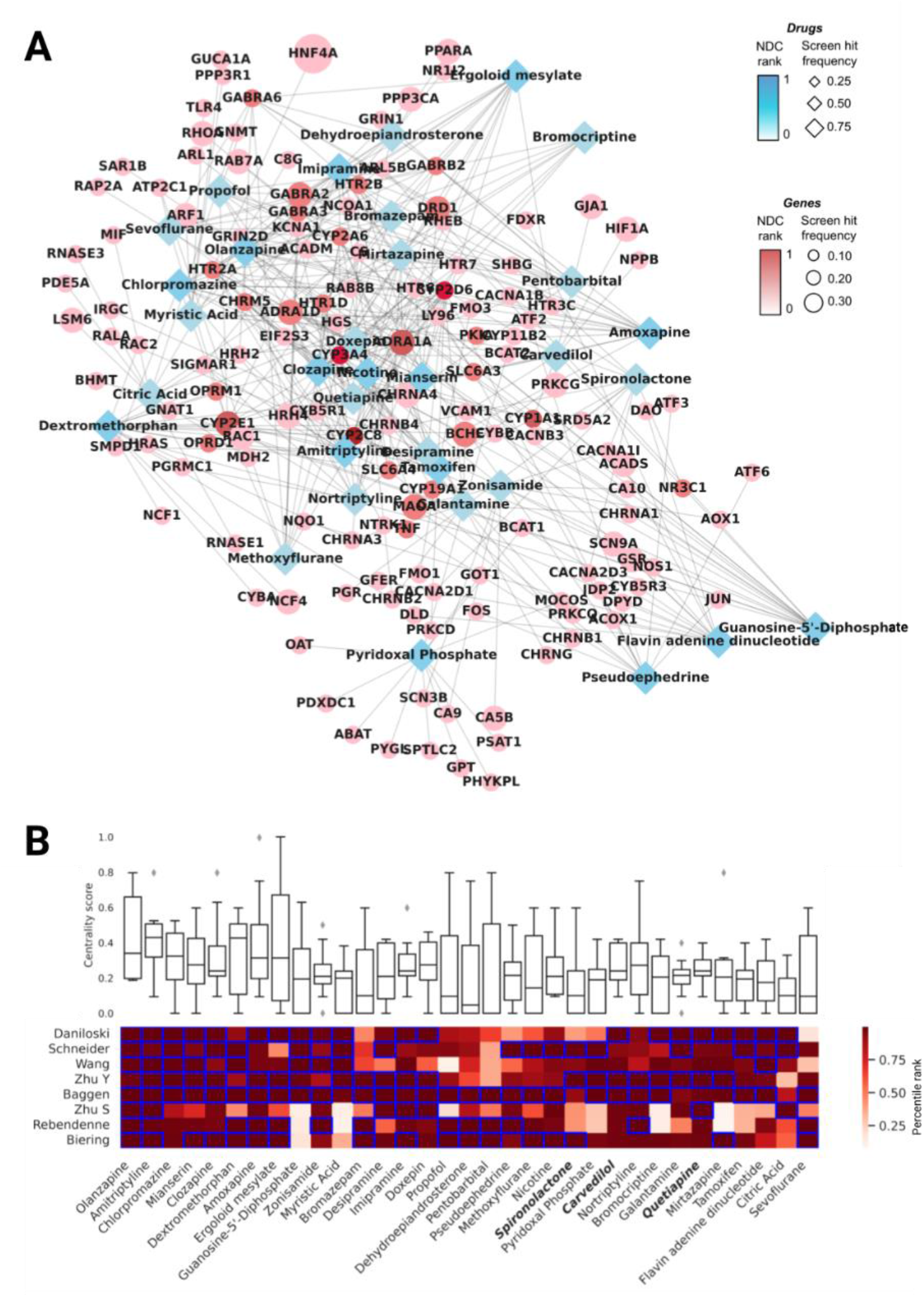
Network-based prioritization of candidate drugs. A. Summary network showing overlap among the individual screen networks. Drug and gene nodes are labeled by average centrality and by the frequency at which they were labeled a hit in individual screens. Psychotropic drugs, particularly tricyclic antidepressants and atypical antipsychotics, show high aggregate centrality, while calcium and potassium channels represent highly targeted entry genes. B. Aggregate ranking of candidate drugs. Bars indicate centrality across all screens. Heat map shows percentile rank in each screen. Blue boxes indicate screens in which the given drug was labeled as significant, and bold text indicates drugs with sufficient cohort numbers for retrospective clinical investigation.

We next performed a propensity-score-matched retrospective clinical analysis to evaluate whether use of candidate drugs was associated with COVID-19 disease severity (**Figure 4A**). We obtained electronic medical records from a large academic hospital system, which yielded 64,349 patient records with a positive COVID-19 test. Of the drugs meeting centrality significance, only three medications had a sufficient treatment cohort size for PSM analysis: carvedilol, quetiapine, and spironolactone (**Figure 4B**). Additionally, we included metformin as a positive control, as it has demonstrated a significant negative association with COVID-19 severity in clinical studies (9, 34–36).

**Figure 4.**
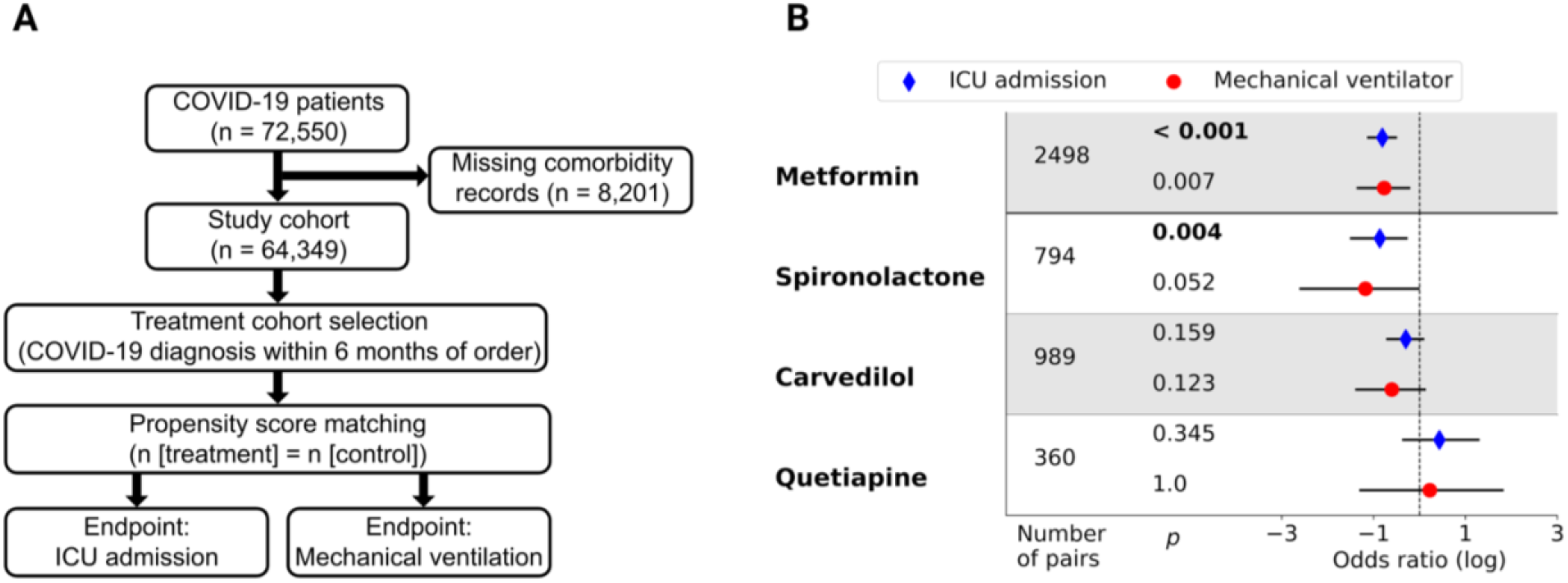
Retrospective clinical validation of candidate drugs. A. Study design. Propensity score matched treatment and control groups were defined for each drug of interest in patients with a COVID-19 diagnosis. Clinical endpoints were ICU admission and mechanical ventilation. B. Cohort sizes, odds ratios, and significance levels for individual drugs and endpoints. Bold text indicates drug-endpoint combinations that were significant after Bonferroni correction (a = 0.05; m = 8). Statistically significant negative associations with ICU admission were observed for both spironolactone and metformin, which was included as a positive control.

We observed a significant negative association between spironolactone use and progression to ICU admission (OR 0.42; CI_95_ 0.22-0.78; p = 0.004). The association between spironolactone use and progression to mechanical ventilator status was also strongly negative, although not statistically significant (OR 0.31; CI_95_ 0.073-1.00; p = 0.052). The positive control, metformin, also exhibited a significant negative association with progression to ICU admission (OR 0.44; CI_95_ 0.32-0.61; p < 0.001) and a nominally significant association with progression to mechanical ventilator status (OR 0.46; CI_95_ 0.25-0.82; p = 0.007). We did not observe a significant association with progression to either ICU admission or mechanical ventilator status for either carvedilol or quetiapine.

Given that spironolactone use was associated with a significant reduction in risk of severe COVID-19 in our cohort analysis, we evaluated whether its mechanism could be mediated by inhibition of viral entry. We performed a SARS-CoV-2 pseudoviral entry assay in a human lung epithelial cell line at varying doses of spironolactone, observing a time- and dose-dependent drug effect on viral entry (**Figure 5A**). We observed an initial peak of viral entry at 4-8 hours after infection, at which time there was a small but significant increase in infection at the highest dose (**Figure 5B**). Following the first peak, there was a larger and sustained decrease in viral infection at higher spironolactone doses, consistent with an overall inhibitory effect on viral entry (**Figure 5C**).

**Figure 5.**
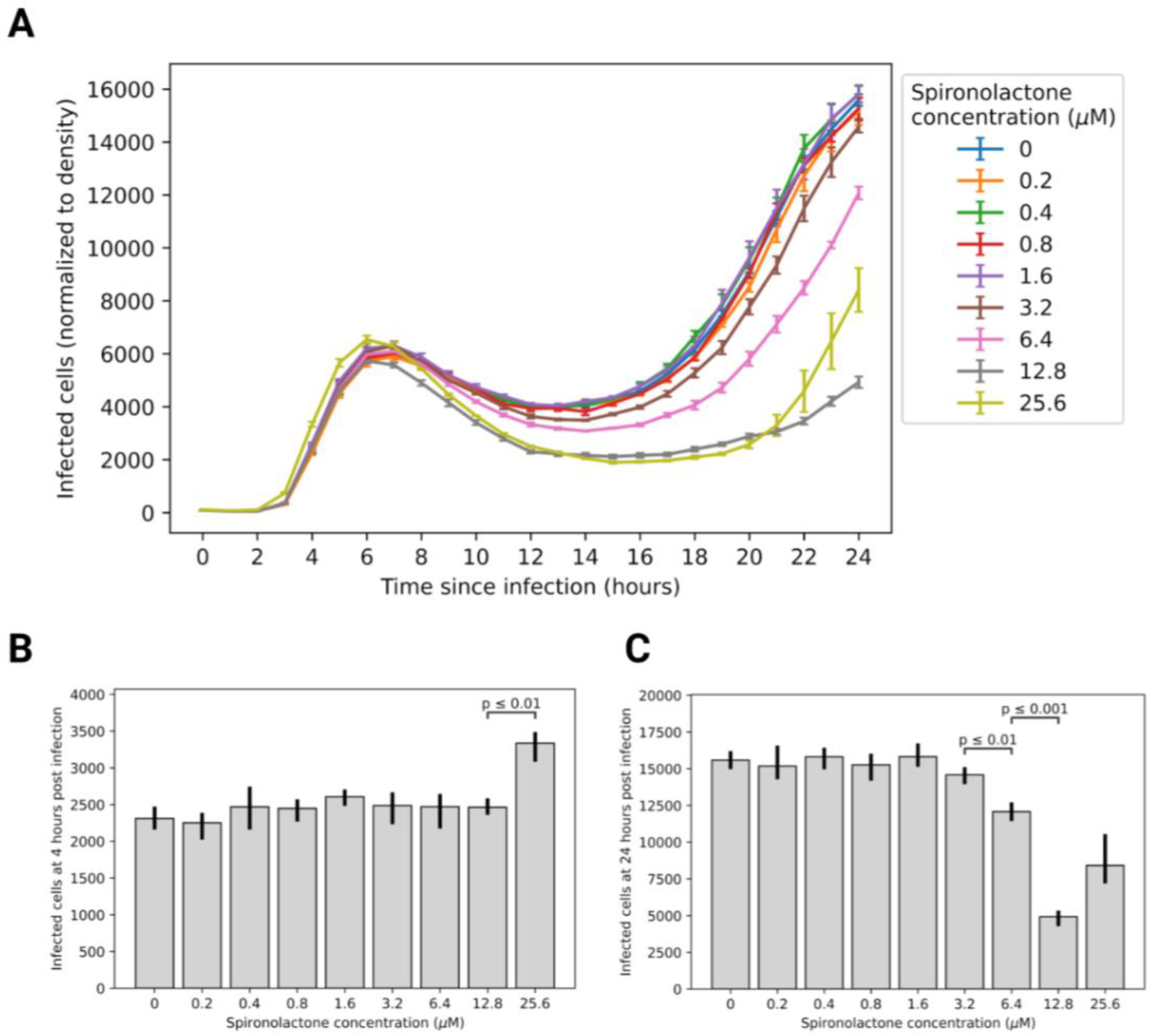
Spironolactone inhibits SARS-CoV-2 pseudoviral entry in human lung cells. A. Infected cell density over time at increasing doses of spironolactone. n = 3 replicates for all time points measured. B. During the initial transitory entry spike at 4 hours, spironolactone was associated with a mild, dose-dependent increase in viral entry, which was significant at the highest dose. C. At 24 hours post infection (steady-state phase), spironolactone conferred a dose-dependent inhibitory effect on viral entry. Brackets show all significant sequential differences.

## DISCUSSION

Our analysis demonstrates that genome-wide CRISPR screens provide a basis for systematic prioritization of drug candidates in COVID-19, many of which are not evident in methods reliant on gene expression studies or association hits alone. We identify three common medications, spironolactone, quetiapine, and carvedilol, as potential modulators of SARS-CoV-2 infection based on their interactions with host entry factors. We perform a propensity-score-matched, retrospective cohort study of clinical outcomes for COVID-19 patients on these medications, showing that spironolactone use is associated with reduced likelihood of ICU admission. We further show that spironolactone inhibits SARS-CoV-2 pseudoviral entry in a human lung epithelial cell line in a dose-dependent manner, providing a potential mechanism for the observed therapeutic effect.

Functional genomic screens enable measurement of the effect of individual host genes on viral entry, but their noise levels and context dependence limit direct clinical applicability. We addressed this barrier by combining data from multiple genome-wide CRISPR screens across several experimental and cellular contexts, inferring drug activity based on abundance of known drug targets among hits, and selecting drugs with high representation among screens for follow-up analysis. Such a prioritization method benefits from direct biological interpretability, whereby top candidate drugs modulate large numbers of entry factors, while enabling higher sensitivity for detecting potential effects.

Pharmacologic antagonists of the renin-angiotensin-aldosterone system (RAAS), such as spironolactone, have been proposed as potential inhibitors of SARS-CoV-2 infection due to their role as indirect modulators of ACE2 expression (37, 38). For instance, spironolactone has been observed to decrease expression of soluble ACE2, which normally promotes viral adhesion, while simultaneously increasing expression of the membrane-associated form (39, 40). Its anti-androgenic action may also decrease expression of TMPRSS2, leading to impaired Spike protein proteolysis and activation (41). Finally, given that COVID-19 is strongly associated with both hypokalemia and hypocalcemia, RAAS antagonists that maintain cation homeostasis, such as spironolactone, may mitigate both ion-channel-mediated viral entry and the clinical sequelae of associated electrolyte imbalances (42, 43). We note that, excluding the cytochrome P450 family, all spironolactone-targeted entry factors in our analysis classify as either androgen signaling proteins (PGR, NR3C1, SRD5A2, and SHBG) or voltage-gated cation channels (CACNB3, CACNA2D1, CACNA1B, CACNA2D3, and CACNA1I), supporting a possible combination of mechanisms. Further studies in relevant cell or tissue contexts could help to elucidate the physiological role of these putative host factors/pathways during viral infection.

Despite a variety of mechanistic hypotheses, few investigations of either clinical efficacy or *in vitro* effect have been performed for spironolactone in COVID-19. A recent interventional study of spironolactone-sitagliptin combination therapy showed statistically non-significant improvements in clinical outcomes, including mortality, ICU admission, intubation rate, and end-organ damage, for patients on spironolactone (44). Another interventional trial showed no significant clinical improvements for COVID-19 patients on potassium canrenoate, a mineralocorticoid receptor antagonist similar to spironolactone (45). However, sample size was limited in both studies, and our retrospective study design enables analysis of a substantially larger treatment cohort.

Our study has several limitations, including the use of a single healthcare system (across 11 hospitals) for clinical analysis, and the use of pseudotyped virus for *in vitro* validation. Further investigations, including well-powered randomized controlled trials, will be necessary to determine the therapeutic role for spironolactone in COVID-19.

## Supporting information

STROBE Checklist

## Data Availability

All data produced in the present study are available upon reasonable request to the authors.

