## Supplementary Information for "Integrative analysis of viral entry networks and clinical outcomes identifies a protective role for spironolactone in severe COVID-19"

p.2. Supplementary Table S1. List of medical comorbidities for cohort analysis.

Supplementary Table S1. **List of medical comorbidities for cohort analysis.**

**Definition:**

AIDS HIV

Cerebrovascular disease

Chronic pulmonary disease

Congestive heart failure

Dementia

Diabetes with chronic complications

Diabetes without chronic complications

Hemiplegia or paraplegia

Malignancy

Metastatic solid tumor

Mild liver disease

Moderate or severe liver disease

Myocardial infarction

Peptic ulcer disease

Peripheral vascular disease

Renal disease

Rheumatic disease
